# A Low-cost, Low-energy Wearable ECG System with Cloud-Based Arrhythmia Detection

**DOI:** 10.1101/2020.08.30.20184770

**Authors:** Nurul Huda, Sadia Khan, Ragib Abid, Samiul Based Shuvo, Mir Maheen Labib, Taufiq Hasan

## Abstract

Continuously monitoring the Electrocardiogram (ECG) is an essential tool for Cardiovascular Disease (CVD) patients. In low-resource countries, the hospitals and health centers do not have adequate ECG systems, and this unavailability exacerbates the patients’ health condition. Lack of skilled physicians, limited availability of continuous ECG monitoring devices, and their high prices, all lead to a higher CVD burden in the developing countries. To address these challenges, we present a low-cost, low-power, and wireless ECG monitoring system with deep learning-based automatic arrhythmia detection. Flexible fabric-based design and the wearable nature of the device enhances the patient’s comfort while facilitating continuous monitoring. An AD8232 chip is used for the ECG Analog Front-End (AFE) with two 450 mi-Ah Li-ion batteries for powering the device. The acquired ECG signal can be transmitted to a smart-device over Bluetooth and subsequently sent to a cloud server for analysis. A 1-D Convolutional Neural Network (CNN) based deep learning model is developed that provides an accuracy of 94.03% in classifying abnormal cardiac rhythm on the MIT-BIH Arrhythmia Database.

**Index Terms:** Wearable ECG, deep learning, arrhythmia detection.

## I. INTRODUCTION

Cardiovascular diseases (CVDs) are considered as the number one cause of death globally (WHO) [1]. Bangladesh has one of the highest prevalence of CVD among the developing world, where 99.6% male and 97.9% female population are exposed to at least one established CVD risk factors [2]. However, the healthcare infrastructure has inadequate resources to diagnose and control the burden of these deadly diseases [3]. In particular, continuous monitoring of the vitals for critical patients is crucial in the case of CVD patients [4].

In Bangladesh, there are approximately 3.05 physicians per 10,000 population, which is extremely low. This lack of skilled healthcare workers makes it difficult to detect and monitor the health status of CVD patients when needed [5], [6]. In addition, number Intensive Care Unit (ICU) beds are also severely inadequate compared to the number of patients [7]. Although portable standard 12-lead ECG systems are available in the market, these are expensive and are not readily available in major public hospitals of Bangladesh, where there is a high burden of patients. Often there are not enough hospital beds for the number of patients admitted, resulting in patients being treated on the floors and hallways [8]. Thus, low-cost and wearable ECG monitoring devices for remote patient monitoring could significantly benefit the hospital systems, especially the cardiology units.

In recent years, many research studies have been conducted on wearable ECG. In [9], a real-time cardiac arrhythmia detection device based on AD8232 and Raspberry Pi (RPI) was proposed. However, detection of the multiple classes ECG arrhythmia was not considered. Another study presented in [10] proposes a wearable device using AD8232, TI C5515 DSP, and RPI-3 to detect only premature ventricular beat (PVC) using wavelet transformation, R-peak identification and template correlation techniques. ECG application expands in the area of Internet of Things (IoT) technology, where the Intel Galileo microcontroller board was used in [11]. In this work, the extracted ECG data were binary classified for abnormal and normal ECG by using a Support Vector Machine (SVM) model. However, the Intel Galileo micro-controller is expensive and not suitable for low-resource settings. In [12], the authors present a detailed review of available wearable ECG devices, including AliveCore, Zio patch, Reka, and Omron heartscan. All of these devices utilize local storage and are too expensive for wide-scale usage in low-income countries.

In this work, we propose a single-lead compact wearable ECG device that costs around $43 and this cost is comparatively lower than other available devices. The device can potentially connect to a computing device (e.g., a smartphone or PC) via Bluetooth, which can transmit the ECG fragments to a cloud-based deep learning server. The ECG signal is automatically analyzed to detect abnormal heart rate and 13 other forms of arrhythmia and alert the physicians accordingly.

## II. Background

Electrocardiogram (abbreviated as ECG or EKG) is a method of measuring the electrical activities of the heart using multiple electrodes placed on the skin of the chest. ECG signal is composed of different segments and intervals, which vary in pathological ways during irregular or abnormal heart rhythms, commonly known as arrhythmia.

An arrhythmia is a group of conditions in which the heart beats irregularly, too fast or too slow. If the heart rate is too fast (exceeding 100 bpm), the condition is known as tachycardia, whereas if it is too low (less than 60 bpm), the condition is known as bradycardia. Arrhythmia may or may not have symptoms, while typical symptoms include palpitation or feeling a pause between heartbeats. Symptoms of a severe case may include light-headedness, shortness of breath, passing out, or chest pain. While most arrhythmias are not harmful, some may cause stroke, heart failure, or even sudden death. An arrhythmia occurs suddenly, and thus continuous monitoring of patients at risk is the only way of early diagnosis and treatment of the resulting health complications [13], [14].

**Fig. 1:**
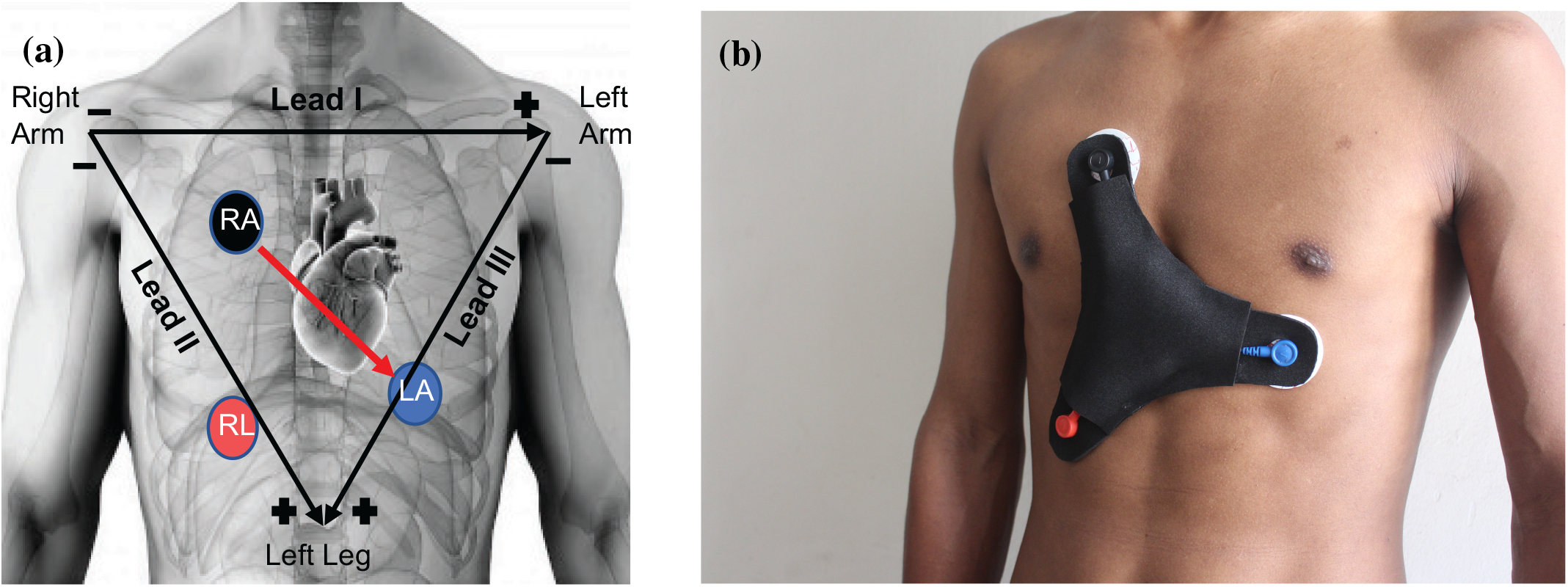
(a) Electrode placement configuration used in the proposed device. Directions of the Lead I-III cardiac vectors are also shown. (b) The proposed wearable device prototype placed on the chest of a healthy volunteer. The Red, Black and Blue electrodes correspond to the Right Leg (RL), Right Arm (RA) and Left Arm (LA) terminals, respectively.

## III. Proposed System Design

In this section, we describe the overall ECG system design in two sub-sections, namely, hardware and software design. The ECG system prototype placed on a test subject is shown in Fig. 2(b). The different components of the design and described in the following sub-sections.

**Fig. 2:**
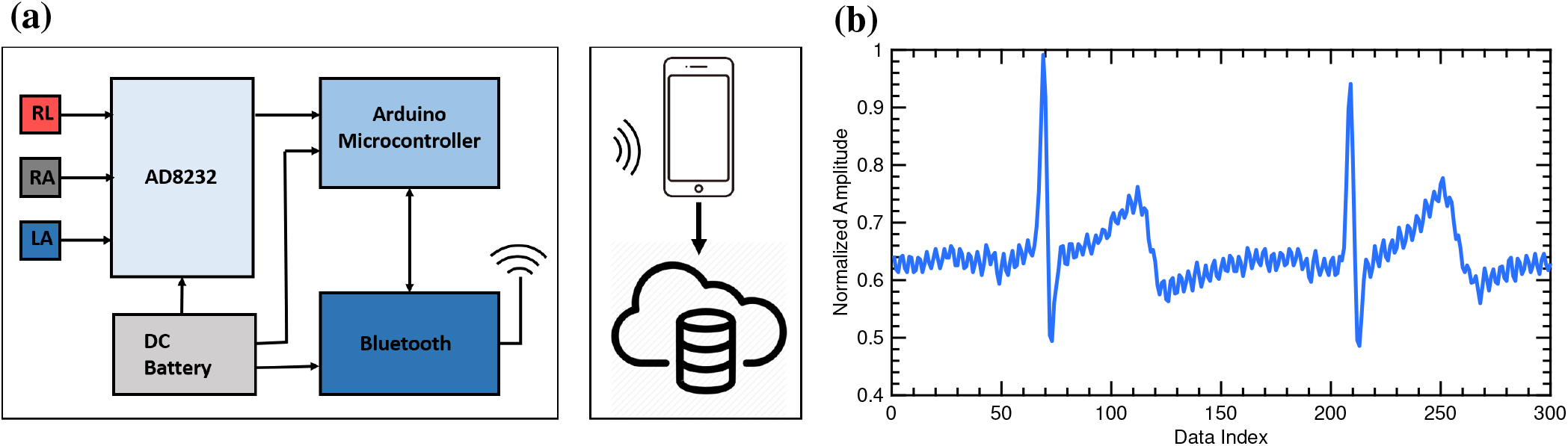
(a) The schematic diagram of the proposed ECG system. The AD8232 is powered up by the DC batteries and receives input from the RA, LA, and RL electrodes. It provides output to the Arduino nano, which is connected with the Bluetooth module to transfer data to a smart device (smartphone or PC). Heart Rate measurement and multiple classes of arrhythmia detection are performed in the cloud-server using deep learning algorithms while the final prediction is transmitted back to the smart-device for alerting the physicians. (b) Raw ECG signal collected from our wearable device.

### A. Hardware Design

The hardware design is performed in four steps. Firstly, the electrode placement and the wearable device dimensions are decided. Next, we considered data acquisition, power management, and data transmission.

1. *Electrode Placement:* Individual heart cells can be regarded as a small electric dipole. The superposition of the electrical activities of all cells of the heart simultaneously can be represented as a single equivalent electrical dipole, known as the cardiac vector. The Einthoven’s triangle, as shown in Fig. 2(a), is an imaginary equilateral triangle defined with the heart at its center and formed by the axes of the three bipolar limb leads, Lead I, II and III. In our design, we aim to focus on measuring the ECG Lead II signal, which is formed by the potential difference between the Left Leg (LL) and Right Arm (RA) electrodes. There are several different electrode placement configurations available for recording the ECG. In this work, we follow the 3-lead configuration system for electrode placement [15]. This electrode configuration is shown in Fig. 2(a) with the electrode locations marked as RA, LA and RL. The Lead II in our configuration is formed between RA (Black) and LA (Blue), which is almost parallel to the standard direction of Lead II cardiac vector according to the Einthoven’s triangle. Therefore, we expect that the heart’s electrical activities along the standard Lead II direction can be observed in the proposed electrode placement configuration.
2. *Data acquisition*: We use the SparkFun Single Lead Heart Rate Monitor AD8232 as the data acquisition device. This device is a complete ECG measurement solution available on a single Integrated Circuit (IC) package consisting of an instrumentation amplifier, a low pass filter, and a leads-off detection with a right leg drive circuit. Signals from the module is then fed to the input of the Analog to Digital Converter (ADC) of an Arduino Nano. The included right leg drive circuit takes in the common-mode voltage from the electrodes by negatively amplifying to drive it back to the body to reduce baseline wandering in the ECG waveform and also eliminates the 50Hz power line interference, if present. Proper lead contact and position is ensured by using floating electrodes and skin-friendly foam pad.
3. *Power management:* For powering the device, two 450mAh Li-ion battery cells and a power management IC is used. For user-friendly application, the system is designed in such a way it can be charged by using a commonly available micro USB cable [16].
4. *Data transmission:* We utilize the HC-05 Bluetooth module for transmitting the ECG data to the smart-device. This device supports a wide range for data transmission and enables free movement of the patient, allowing distant monitoring by the physician. The final device specifications are summarized in Table I.

**TABLE I:** Device specification

| Parameter | Specification |
| --- | --- |
| Dimension | 4 inch × 4 inch |
| Electrode type | Floating electrode |
| Battery Type | Rechargeable Li-ion |
| Battery Capacity | 900mAh |
| Wireless Transmission | Bluetooth Module - HC-05 |
| Range | 10m |

### B. Software Design

In this section, we describe the cloud-based deep learning server for arrhythmia detection.

1. *Training dataset:* Our arrhythmia detection algorithm is based on a deep CNN architecture based on [17], [18]. The MIT-BIH arrhythmia dataset obtained from Physionet is used to train the CNN model. This dataset contains ECG signals from 47 subjects recorded for 24-hours [19]. Among them, we select data from 33 subjects for classifying 14 classes of rhythm. In order to test the models, 30% of the data is heldout, while the remaining data is used for training. Since our wearable ECG is designed to capture the ECG Lead II, we use only this lead from the MIT BIH dataset.
2. *Pre-processing:* We used an anti-aliasing filter and a low-pass filter to remove unwanted noise and perform R-peak detection using the Pan-Tompkins algorithm [20]. The duration of the ECG signal from a single subject is 30.05 minutes, from which we select segments consisting of 3 R peaks and perform zero-padding to ensure a constant length. The segments are labeled with the corresponding annotation provided. These segments of rhythm are then used for training the CNN model for arrhythmia detection [17], [18].
3. *CNN model architecture:* The CNN model architecture is illustrated in Fig. 3. It consists of 1-D convolution, max-pooling, batch normalization, and dropout layers. The flattened layer output is passed through a fully connected layer (with dropout) and a second fully connected dense layer (with dropout). Finally, a SoftMax layer with 14 outputs is used for arrhythmia classification. The 14 types of rhythm that is detected by the CNN model is provided in Table II. A balanced mini-batch training scheme is used to address the data imbalance issue where every 14 classes of data are re-sampled to provide a balanced input data of 2000 segments. We obtain a training set accuracy of about 97.53% and validation set accuracy of about 94.13%. In addition to detecting abnormal ECG patterns, the heart rate is also calculated from the R-R intervals.
4. *Real-time processing:* Data from the wearable ECG system is received in the smart-device via the Bluetooth module. This segment of data is then sent to the smart-device, which transmits the data to the server for further processing, segmentation, and testing. Pre-processing and R-peak detection are performed in the same manner as the training data. The detected R-R intervals used to compute the heart rate and each rhythm, which consists of 4 R peaks, is then passed to the CNN model to detect other types of abnormal heart rhythm as summarized in Table II. The test result is transmitted back to the display of the smart-device. If the application determines three consecutive abnormal rhythms, it provides a warning alarm to notify the physician. The current version of the system is implemented on a desktop PC in place of a smart-phone.

**TABLE II:** Classes of ECG Rhythms

| Symbol | Rhythm Description | Label |
| --- | --- | --- |
| N | Normal Sinus Rhythm | 0 |
| AB | Atrial Bigeminy | 1 |
| AFIB | Atrial fibrillation | 2 |
| AFL | Atrial flutter | 3 |
| B | Ventricular bigeminy | 4 |
| BII | 2° heart block | 5 |
| T | Ventricular trigeminy | 6 |
| VFL | Ventricular flutter | 7 |
| VT | Ventricular tachycardia | 8 |
| SVTA | Supraventricular tachyarrhythmia | 9 |
| IVR | Idioventricular rhythm | 10 |
| P | Paced rhythm | 11 |
| SBR | Sinus bradycardia | 12 |
| NOD | Nodal (A-V junctional) rhythm | 13 |

**Fig. 3:**
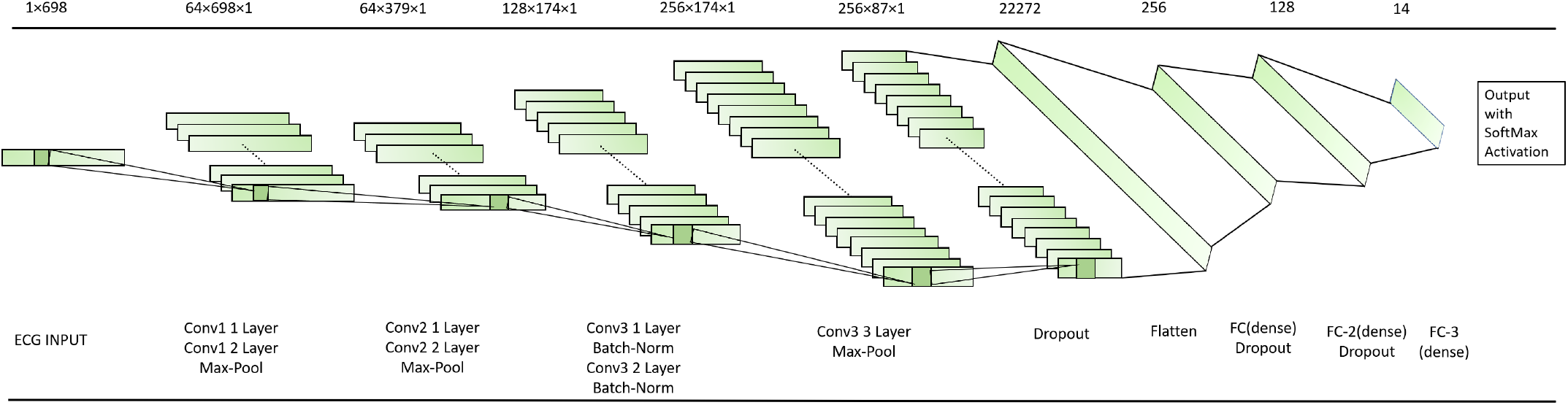
The 1D-CNN model architecture used for arrhythmia detection.

## IV. Experimental Evaluation

The epoch-wise accuracy and loss for our model during training and validation are shown in Fig. 4 and 5, respectively. On the MIH-BIH held-out test set, the model provides an averaged accuracy of 94.03% over the 14 classes. The average sensitivity, specificity and F-1 scores of the system are found to be 0.940 (±0.087), 0.995 (±0.005), and 0.939 (±0.0689), respectively.

**Fig. 4:**
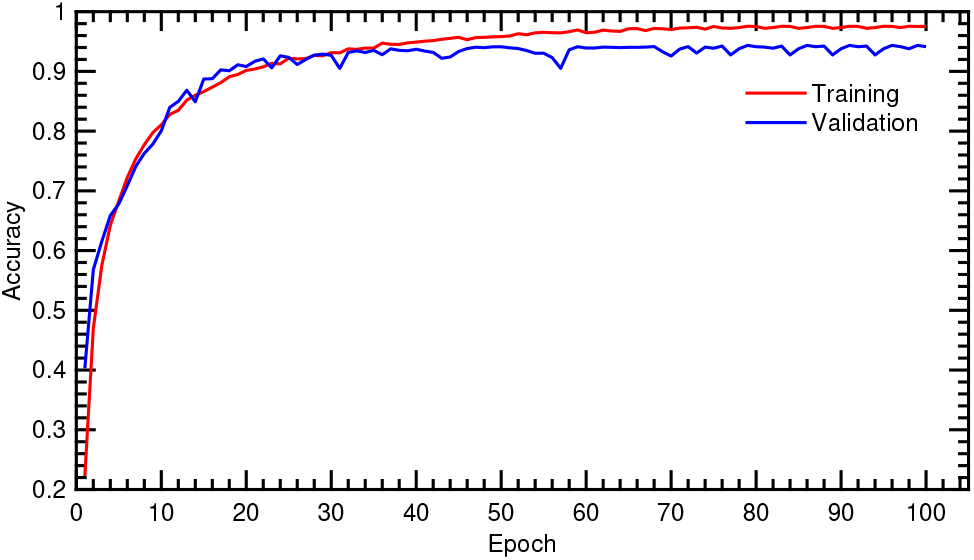
Epoch vs accuracy of the CNN during training and validation.

**Fig. 5:**
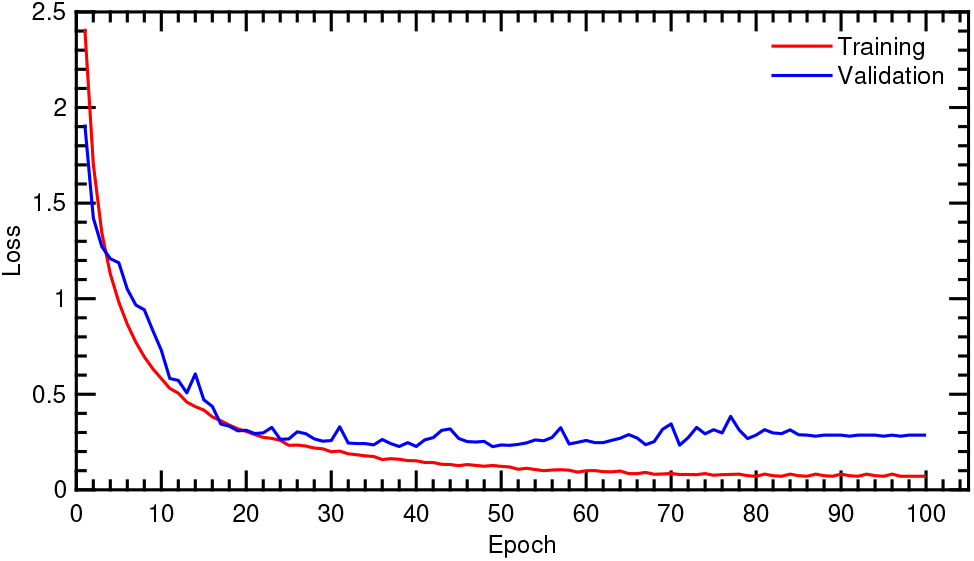
Epoch vs loss of the CNN during training and validation.

Experimental evaluation on CVD patients is not yet performed using this device as we are still in the process of gaining ethical clearance and approval from an Institutional Review Board (IRB). However, in informal tests with five healthy subjects, four of the subjects resulted in a correct diagnosis of ”normal”. Further evaluations and extensive data collection from CVD patients are required to fine-tuning the CNN model and improve its performance on real-world data.

## Conclusions

In this work, we have developed a low-cost wearable ECG system with the low-power requirement. The device is capable of connecting to a smart device (e.g., a PC or a smart-phone) via Bluetooth and automatically assess different types of arrhythmia. A flexible fabric-based wearable construction was used for the device that is designed to capture the ECG Lead II. The ECG instrumentation was based on an AD8232 neat little chip powered by two 450 mi-Ah Li-ion batteries. The back-end classifier uses a one-dimensional CNN model trained on the MIT-BIH dataset to detect 14 different types of arrhythmia, including Atrial Fibrillation (AFIB) and Ventricular Flutter (VFL). The system obtained an overall accuracy of 94.03%.

## Data Availability

Data used in this research are open-sourced. References are provided in the manuscript.

